# Benchmarking Large Language AI Models Against Human Doctors Using an Emergency Medicine Simulation Approach

**DOI:** 10.64898/2026.11.09.26367590

**Authors:** Masoud Shahabian, Ali Hadizadeh Esfahani, Mehdi Ahmadi, Somayeh Rajabzadeh, Mohamad Hasan Givechin Ghamsari, Arash Tabaraee, Seyed Zia Hejripour Rafsanjani

## Abstract

**Background:** Proper adoption of AI in emergency medicine necessitate thorough and rigorous appraisal. Classic benchmarks, which rely heavily on basic question-and-answer formats, though dominating the benchmarking field of LLM usage in medicine, have proven inadequate in assessing the full spectrum of skills incumbent upon an agent in healthcare.

**Methods:** We propose an intricate benchmarking scheme in which the skill set required of an emergency physician is tested and quantified. Our benchmarking structure is 3-fold: skills such as history taking, diagnostic accuracy, treatment plan, and empathy are evaluated by an emergency medicine expert, while the same expertise is scored by an intern from the perspective of a patient, through what we call a simulated emergency room visit. A comprehensive written examination is also utilized to measure general emergency medicine knowledge.

**Results:** Eight LLMs were benchmarked against each other and in contrast to nine emergency medical doctors. Our findings indicate that the AI models significantly outperformed human physicians across several key domains, including history taking, investigation, diagnostic accuracy, and treatment planning. Furthermore, the models demonstrated superior performance in comprehensibility, credibility, and empathy. When comparing AI models against each other, performance advantages were marginal during simulated emergency room visits and were mostly pronounced during the written examination.

**Conclusion:** AI models demonstrate a clear advantage over human physicians across a diverse range of emergency medicine competencies. The proposed benchmarking scheme provides a comprehensive metric for a thorough evaluation of medical skill set, furthering validation, adaptation, and utilization of AI in emergency medicine.

## Introduction

### Background

Diagnostic error, i.e., a clinician’s oversight of a diagnosis or failure to reach a correct one in a timely manner, is a pervasive global occurrence and among the prime causes of preventable harm in the healthcare system [1, 2].

In emergency medicine (EM), rapid and accurate information gathering, decision-making and treatment planning are of paramount importance. However, the heuristics of emergency medical doctors are constantly under disruptive pressure from the high-stakes nature of EM; doctors have to make multitudinous life-saving decisions under time pressures and in disorderly situations. These factors, among others, hinder the diagnostic skills of clinicians, prompting research in methods tackling these factors and facilitating accurate diagnosis in EM [3].

Alongside the explosive advancements of Artificial Intelligence (AI) and the widespread utilization in different fields, it has been incorporated in EM as well. Specific sub-classes of AI, e.g., Large Language Models (LLMs), are suited for tasks such as gathering information and clinical decision-making. By integrating LLM assistance, one could subdue the eroding factors of working in EM, culminating in the reduction of diagnostic errors [3, 4]. LLMs can streamline history and information taking from the patient, which speeds up the process and reduces the cognitive load and the chance of cognitive fatigue [5, 6]. They can also support the clinician through the decision-making process, increasing accuracy and patient throughput, decreasing the chance of cognitive bias, and overall, reducing the probability of a diagnostic mistake [7, 8]. Moreover, LLMs are shown to outperform physicians in diagnostic reasoning depending on the context, making them ideal to be part of an AI-based support system [9].

### Importance

LLM utilization in EM requires thoughtful design, meticulous validation, and careful attention to ethical and practical considerations; hasty adoption of LLMs could ironically result in an increase in errors in ED, endangering patients’ health, diminishing trust in AI, and therefore hampering the global adoption of such tools in EDs [3, 10].

Innumerable research projects have been dedicated to evaluating AI applicability and safety in the medical context. Since LLMs’ medical theoretical knowledge forms the foundation for their medical competence, initial efforts focused on compiling multiple-choice medical questions [11-14]. On top of evaluating medical knowledge, attempts to appraise clinical reasoning and decision-making skills were made through benchmarking datasets [15-17]. There are disputes about these types of benchmarks, however, arguing that simple question-and-answer approaches don’t represent the clinical utility of AI completely, nor do the more complex and synthetic computational simulations of clinical practice [18, 19]. Validating and implementing AI as a safe and accepted EM tool necessitates thoughtful design and thorough investigation schemes, by involving EM clinicians and patients as stakeholders in the whole benchmarking process [3].

### Goals of This Investigation

In this paper, we propose an extensive evaluation process to measure an LLM’s medical capabilities in the ED. Our benchmarking approach is based on simulating EM; by putting an LLM in simulated emergency scenarios as an EM clinician, not only can the medical knowledge and diagnostic proficiencies be analyzed, but we can also properly evaluate the other necessary skills, such as history-taking or empathy [20]. To contrast against the common benchmarking approaches, we also design a written exam to evaluate EM knowledge and ascertain the competency of LLMs and EM doctors. Overall, our work provides a comprehensive insight into different types of LLMs’ performance capabilities for the ED (Fig 1).

**Fig 1.**
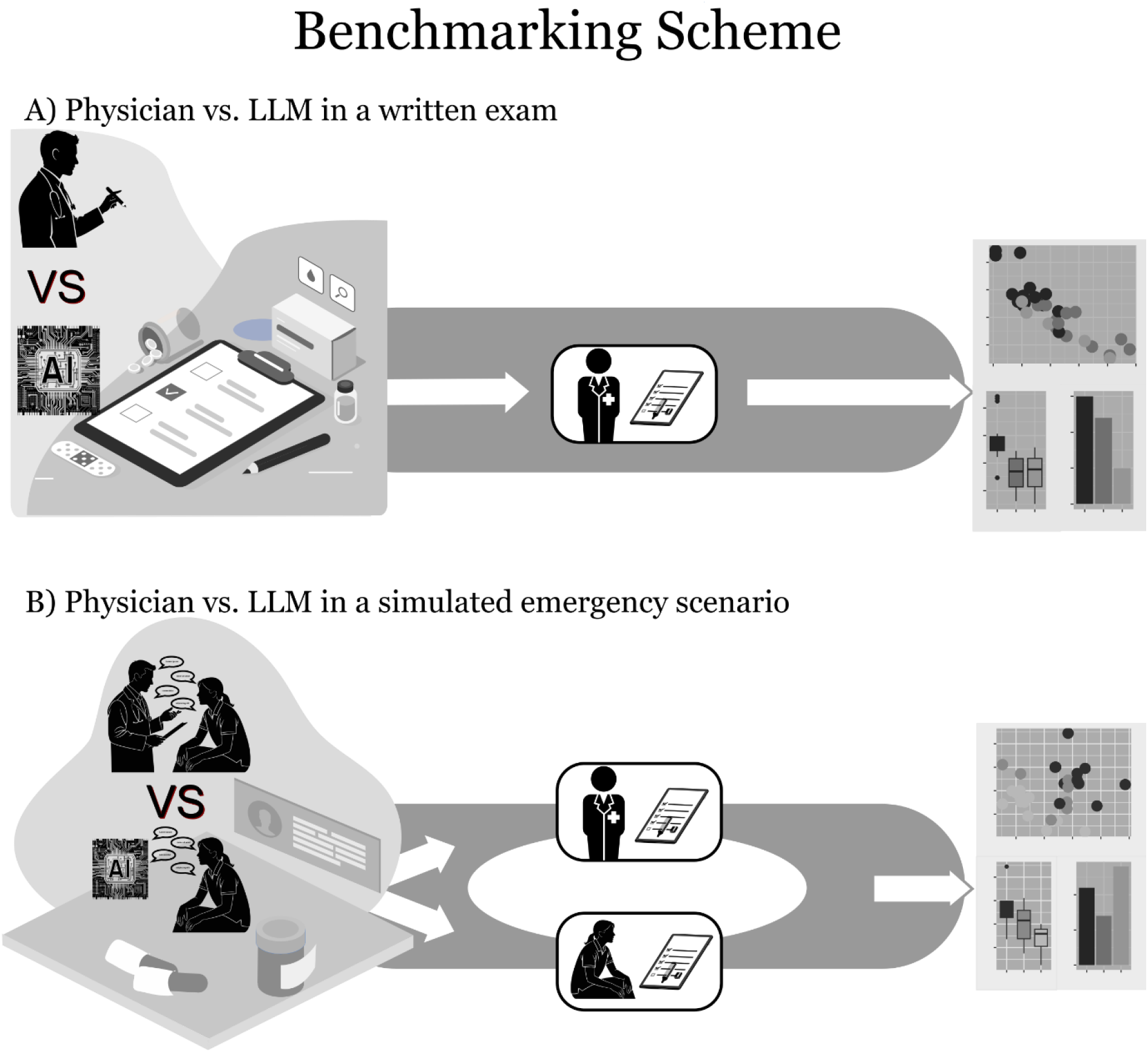
Emergency Medicine Benchmarking of AI. **A**. Both LLMs and human physicians take a written exam comprising of 27 emergency medicine cases, each followed by two to three questions. The answers are corrected and scored by an emergency medicine expert. **B**. Interns are trained on three different emergency cases to act as a simulated patient. They are interviewed by both LLMs and human physicians. Each interview is scored by both the emergency medicine expert and the interviewed intern.

## Methods

### Study Design

To establish a baseline to compare the LLMs against, we employed nine EM physicians with different levels of expertise: three residents in early years of residency, three last-year residents, and three emergency medicine specialists. The selected physicians were compared against eight different LLMs, namely ChatGPT 5.6 Sol, Claude 4.8 Opus, DeepSeek R1, DeepSeek V4 Pro, Gemini 3.1 Pro, O3, Qwen 3.7 Max (Fast), and Qwen 3.7 Max (Thinking), all accessed in June 2026.

To achieve an adequate emulation, we utilized real emergency cases that have occurred in the ED. Making use of the book “100 Cases in Emergency Medicine and Critical Care” [21], we randomly^1^ chose thirty emergency cases, three cases for EM simulation (one per clinician in the three-person groups of residents and specialists) and twenty-seven for the written exam.

### Written Exam

The twenty-seven randomly choses cases, each followed by two to three questions, were provided to the eight LLMs, and their corresponding answers were retrieved. For testing the human doctors, each clinician was tested with 9 out of 27 cases, in a way that each group (residents, last-year residents, and specialists) would be tested with all 27 cases in the exam questionnaire. Written answers from clinicians were also retrieved. Both the answers of the LLMs and clinicians were given to the emergency medicine specialist expert to score on aspects such as diagnosis, investigation, and treatment (Fig 1A).

### EM Simulation

For each physician in the experiment, we recruited an intern to represent an emulated patient in an ED case. After being informed and instructed about the case, each intern, acting as an emergency patient, was interviewed by one human physician and eight different LLMs. All the interviews were in Farsi language and performed through a mediator, so that the interns wouldn’t know if they were being interviewed by a human doctor or AI. All the clinicians, interns, and LLMs were instructed beforehand on how the experiment was performed and what was expected from them. After each interview, the intern was provided with a feedback form to rate the interview from different aspects, such as diagnosis, concern reduction, or treatment plan, on a 5-point Likert scale. In addition, the written records of the interviews were given to an emergency medicine specialist expert to rate the performance of clinicians and LLMs on different criteria, namely history taking, diagnostic accuracy, treatment plan, and the level of empathy. Names were removed from the interview records; hence, the expert was blinded to the identity of the interviewer, human or LLM (Fig 1B).

### Data Analysis

All the gathered data in this work is in the format of a 5-point Likert scale, which means that in this work we deal with ordinal data. Therefore, we generally utilized nonparametric methods for processing and analyzing the data. We used the Mann-Whitney U test to compare Likert scores from two different groups. For comparing more than two populations, we employed the Kruskal-Wallis test. To perform post-hoc power analysis for the aforementioned tests, we used Monte Carlo simulation to estimate the empirical power of Mann-Whitney U and Kruskal-Wallis tests. To reduce the dimensions and extract the underlying factors from the multi-dimensional Likert data, we used factor analysis based on polychoric correlation. Detailed explanation about the methods, used packages, the software and the code that performed the analysis is provided in the supplements (S1).

## Results

We used the following nomenclature to distinguish the results of different types:

I. *Simulated Doctor Visit* refers to results that are acquired from EM simulation, resulting from the EM expert scoring the interview records.
II. *Simulated Patient Experience* designates the results from the feedback form received from interns who were acting as simulated patients in EM simulation.
III. *Written Exam* is the result of the EM expert scoring the written answers of clinicians and AI to the written exam.

Firstly, results from LLMs and physicians were plotted separately as horizontal stacked bar charts, also known as Likert plots (Fig 2). Scores from each category, e.g., history taking, empathy, etc., were compared between the human physicians and the LLMs using a directional Mann–Whitney *U* test, testing whether LLMs’ scores are significantly higher than the physicians’, and the resulted *p* values are reported in the central column of Figure 2.

**Fig 2.**
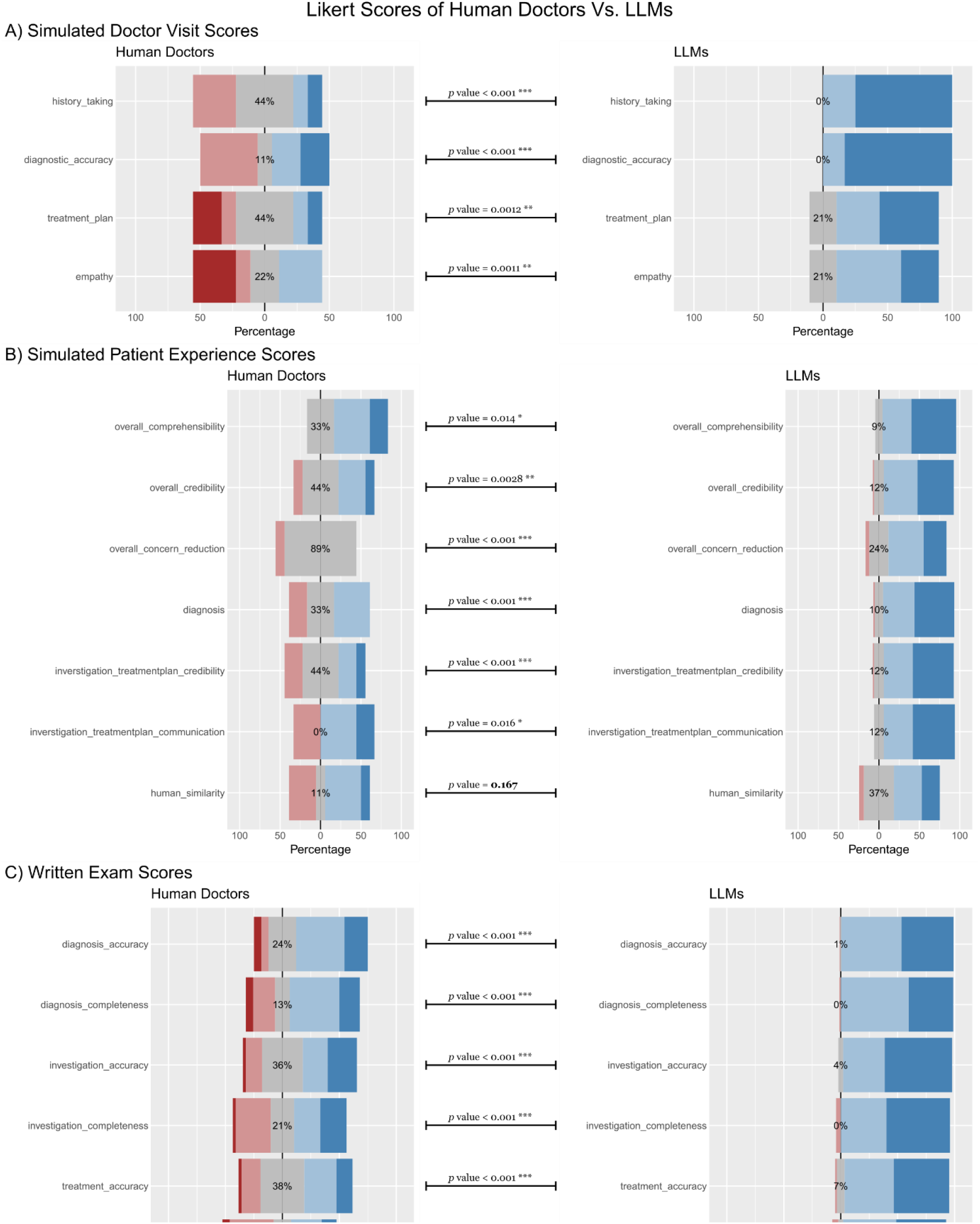
AI vs. Human Physician Likert Scores from Emergency Medicine Benchmarking. In each panels, human physician Likert scores are plotted on the right, LLM scores on the left, and the results from statistical tests are reported in the middle. **A**. Likert scores that are acquired from EM simulation, resulting from the EM expert scoring the interview records. **B**. Likert scores from the feedback form received from interns who were acting as simulated patients in EM simulation. **C**. Likert scores from the EM expert scoring the written exam.

As is evident from the charts and the results of the statistical tests, LLMs scored predominantly higher in almost all categories. In every single category the written exam, LLMs scored significantly higher than physicians, with the *p* values suggesting an extremely significant difference, i.e., *p* < 0.001, marked by three asterisks (Fig 2C), and post-hoc power analysis estimating empirical power at ∼1. The same can be said about values from the simulated doctor visit, with *p* values pointing toward a highly significant advantage, i.e., 0.001 ≤ p < 0.01 marked by two asterisks, or an extreme advantage for LLMs (Fig 2A), with estimated empirical power of ∼0.97. The only group of results in which LLMs didn’t score significantly higher in all categories was simulated patients’ opinions; while LLM results were extremely significant in some categories, such as diagnosis or concern reduction, they were only slightly significant (0.01 ≤ p < 0.05, marked by one asterisk) in others, such as communication or comprehensibility (Fig 2B), and the estimated empirical power was ∼0.83.

When acquiring simulated patients’ feedback after each visit, we queried about their opinion regarding the similarity of the doctor, human or AI, to a human. Patients graded this similarity on a 5-point Likert scale, with 1 meaning a very artificial and ‘robotic’ interaction and 5 referring to a very natural and human-like interaction. Since LLMs were instructed not to reveal that they are AI, and the patients were blinded to the identity of their visiting doctors, this score worked as a Turing test [22]. Ironically, from patients’ point of view, the only category with non-significant difference between human physicians and LLMs was human similarity (Fig 2B).

For a deep, granular look into the performance of each LLM and comparing them against each other, it is possible to plot the Likert scores for every single LLM in each category separately (Supp. Fig 1A). However, having multiple groups of results, each one with multiple subgroups and categories, represents a complex data structure which makes it difficult to properly quantify the differences among LLMs. Using machine learning and data mining approaches, we aimed to simplify the data structure, extract the relevant information, and calculate the performance differences. Firstly, we used factor analysis to extract the underlying factors in the data, so that data dimensions are reduced while relevant information is retained. Scree plots show that there is one underlying factor in simulated doctor visit scores, one in simulated patient experience scores, and three in written exam resulting data (Fig S3). We used factor analysis to extract the underlying factors in doctor visit and patient experience data sets. As for written exam data, we averaged over diagnosis, investigation, and treatment categories to have three interpretable dimensions of data. We then used linear regression modeling on this 5-dimensional data, 5-dimensions being the dependent variables and the type of LLM used as independent or

explanatory variable. As is common in the field, we used the performance of ChatGPT in our experiment as the reference and compared the performance of other LLMs against it. In the fitted linear regression models, each LLM in our work is represented by one term in the models, and the corresponding coefficients are plotted in a forest plot and an accompanying table (Fig 3). Each dot in the forest plot represents the average performance difference of an LLM compared to ChatGPT on a Likert scale, and the whiskers around the dots show the extent of the 95% confidence interval. For example, the value for Gemini in the simulated doctor visit model is -0.78, which means Gemini scored, on average, 0.78 worse than ChatGPT in simulated doctor visit grades.

**Fig 3.**
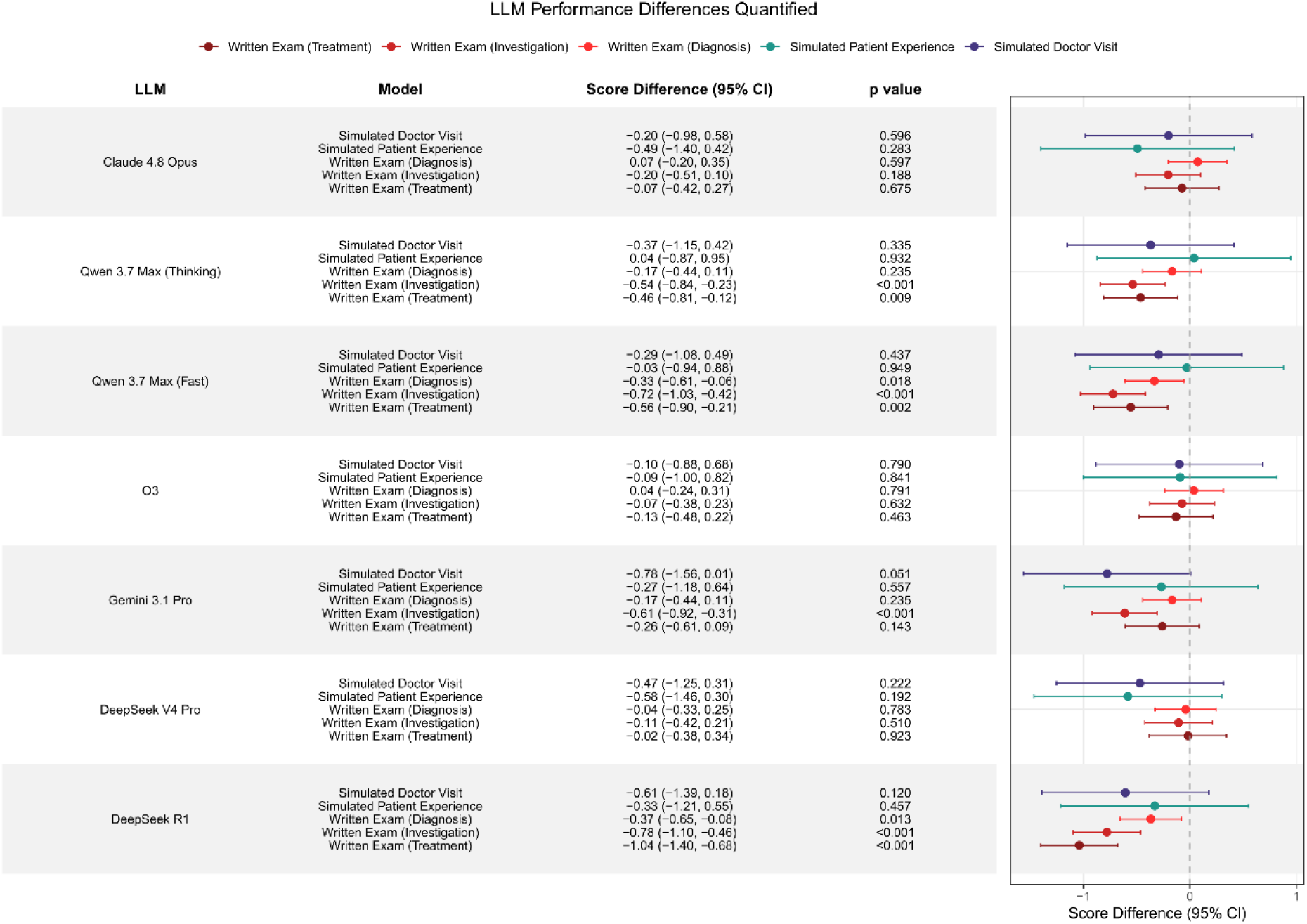
Comparison of Different LLMs Against ChatGPT 5.6 Sol. Linear regression is used to model dimensionally-reduced Likert score results, dimension reduction done using factor analysis. The five models are ‘Simulated Doctor Visit’, ‘Simulated Patient Experience’, ‘Written exam (Diagnosis)’, ‘Written exam (Investigation)’, and ‘Written exam (Treatment)’. Each dot in the forest plot represents the average performance difference of an LLM compared to ChatGPT on a Likert scale, and the whiskers around the dots show the extent of the 95% confidence interval. For example, the value for Gemini in the simulated doctor visit model is -0.78, which means Gemini scored, on average, 0.78 worse than ChatGPT in simulated doctor visit grades.

The values from Figure 3 can paint a proper picture of how the LLMs were measured against each other. As mentioned, values in the figure represent the performance difference relative to ChatGPT as the reference. In addition, almost all values plotted are negative, meaning that other LLMs have performed worse in almost every aspect. Hence, ChatGPT 5.6 Sol is the most competent LLM in EM based on our benchmarking. Three other LLMs could score higher in at least one category: Claude 4.8 Opus, O3, and Qwen 3.7 max thinking, with the latter scoring slightly higher in simulated patient experience score and the other two scoring slightly better in written exam (diagnosis), while scoring worse in the other four categories. On the other side of the spectrum, DeepSeek R1 is located, with all three written exam criteria (diagnosis, investigation, and treatment) scoring significantly worse than ChatGPT. The other three LLMs, namely DeepSeek V4 Pro, Gemini 3.1 Pro, and Qwen 3.7 max fast, are placed somewhere in the middle. Qwen 3.7 max fast performed similarly, but slightly worse to its thinking counterpart. DeepSeek V4 Pro performance was comparable to ChatGPT in written exam values, but worse in simulated doctor visit and patient experience scores. And finally, Gemini 3.1 Pro is placed worse than DeepSeek V4 Pro but better than R1, with only performing significantly worse in one criterion: written exam (treatment).

## Discussions

AI systems can be trained to identify patterns within convoluted datasets, enabling prediction of specific outcomes [4]. In the context of EM, such methods can facilitate triage and provide diagnostic and treatment support, among other tasks [23, 24].

Comprehensive benchmarks have been implemented to evaluate AI in the context of EM. They range from simple approaches that assess a singular skill to holistic frameworks that quantify the competence of AI from multiple angles [11, 25]. Most usually took the same approach: compiling a compendium of questions and tasks, aiming to make each dataset larger than the state of the art, with the goal of covering as many necessary EM skills as possible while circumventing challenges such as data leakage [26, 27]. We took a different approach, however; rather than separately and explicitly testing AI for each specific skill, we aimed to simulate EM and place AI in the simulation. This way, all necessary skills are intrinsically tested, and the results more accurately reflect real-world performance.

The results demonstrated a clear advantage for the tested AI compared to the human physicians. LLMs scored significantly higher in skills such as history taking, investigation, diagnosis, treatment plan, comprehensibility, credibility, concern reduction, and empathy. Unlike the clear contrast between the performances of LLMs and human physicians, the medical expertise among different LLMs was not distinguishable as clearly. In general, some AI models scored higher than others, but the differences were minor and non-significant. The only clear distinction was observed in the written exam, in which ChatGPT, O3, Claude, and DeepSeek V4 performed better than Qwen and Gemini, and DeepSeek R1 performed almost universally worse than others.

Findings were consistent between EM simulation and written exam results. However, there were evaluation aspects that only became evident in the simulation scenarios. Interview length was a variable factor for instance: on average, Gemini reached a diagnosis after asking 6 questions, Claude after around 9, and ChatGPT or O3 after about 12 questions. Since there were no significant differences among LLMs in their history taking and diagnosis scores (Fig S1A-I), the higher interview pace suggests a nimbler conduct. In the high-stakes, chaotic environment of EM, agile performance is a great asset. Additionally, quicker diagnosis signifies far fewer tokens spent. At scale, such disparity can translate to differences in cost and the required resources.

Since the medical competence benchmarking was done in Farsi, it simultaneously operated as a linguistic bias test. English is the most common language in LLM training, resulting in a bias toward this language [28-30]. Analyzing how an AI handles a less common language provides a window into how language bias can affect the interaction quality of LLMs. In our experiments, LLMs handled Farsi communication well for the most part, as reflected in the high comprehensibility scores by the simulated patients (Fig 2). It seemed that most LLMs performed in their original language (English or Chinese, depending on the origin of the LLM), and translated the message in either the last or first stage. Therefore, the quality of the conversation was dependent on the quality of the translation. In rare cases, LLMs produced calques, reducing the quality of the translation. For example, in one instance, in an attempt to provide reassurance, Claude literally translated the term “you are in good hands” to Farsi, which became meaningless and resulted in patient confusion.

Linguistic bias can be classified as a sub-category of cultural bias. On a broader scale, training a language model predominantly in one language and attempting to interact with humans in another can hinder the quality of communication due to the inherent cultural discrepancy between the languages. An example of such bias was the frequent attempts by one of the LLMs to reassure or empathize, which made a simulated patient feel uneasy and doubtful. He was used to interacting with direct and to-the-point doctors, and his interaction with the LLM felt dishonest and insincere. This sentiment was supported by some others during our experiments, leading us to believe that developing one model and using translation to utilize it in different countries might result in impaired communications. If the aim is to deploy a medical AI system on a commercial scale that can properly interact with the patients, adaptations based on the target country are needed.

Since their inception, LLMs have been marred by hallucinations. With each new iteration of an LLM, measures have been taken to reduce the rate of hallucinations [31]. Considering all the different LLMs that have been thoroughly tested in the medical context, this work provided an apt testing environment for hallucination occurrence. During the patient interviews in the course of the EM simulation, no hallucinations were detected. We speculate this is because most of the LLMs used in this work are rather new, and have modern mitigation strategies to prevent hallucinations. The same was true during the written exam benchmarking, with the exception of Claude 4.8 Opus; when the questions in the written exam were given to Claude all together, it answered the first few, then started hallucinating new questions answering its own made-up questions. Multiple tries resulted in the same outcome, forcing us to divide the written exam into smaller batches of questions and gather the answers batch-wise. This shows that while the hallucination rate has been reduced drastically in newer LLMs, they still occur. In medicine, these occurrences can have drastic consequences, necessitating further improvement in hallucination prevention.

In this work, we have demonstrated how novel benchmarking approaches can provide better insights into AI performance in distinctive applications. The added insight comes at a price, however, warranting extra time and resources compared to automated Q&A-based methods. A potential countermeasure for the costly steps would be to partially automate the framework. Admittedly, major ethical, legal, and regulatory challenges for the fully automated benchmarks persist [1]. Yet, approaches such as human-in-the-loop artificial intelligence solutions can mitigate these challenges by balancing automation efficiency with human oversight and improve accuracy beyond unassisted human or AI performance alone [32]. Predictive models can also circumvent some steps which are time- and resource-costly. A solution to use publicly available benchmarking results to predict AI competence in EM is explored in the supplements (S4 and Fig S2).

AI advancements were astonishing in the past few years. In this work, AI could outperform human physicians in almost every metric (Fig 2). Even in a metric such as the *human similarity*, AI performed quite well, indicating that patients found AI and human doctors indistinguishable. The widespread utilization of such competent medical tools might be enticing to the stakeholders. However, any mishap tracing back to impetuous AI employment can have devastating repercussions for the AI acclimation in medicine [10]. Though still limited, negative perception is growing against AI in medicine, due to the usage of imperfect AI programs by some U.S. health insurers and the subsequent sinister effect on patients’ medical care [33, 34]. Decision-makers shall tread carefully, establish stringent regulations, and employ thorough verification if they want to avoid similar unfortunate incidents in emergency medicine.

## Data Availability

All data produced are available online at https://github.com/Ali-esfa/LLM-in-EM

## Footnotes

1 Using NumPy library in Python

