## Supplementary Material for "Benchmarking Large Language AI Models Against Human Doctors Using an Emergency Medicine Simulation Approach"

#### Supplementary Methods

##### S1. Data Analysis

As mentioned in the main text, all the gathered data in this work is in the format of a 5-point Likert scale, which means that in this work we deal with ordinal data, i.e., categorical data where variables have a natural order. Therefore, we generally utilized nonparametric methods for processing and analyzing the data. The Mann-Whitney U test, also known as the Wilcoxon rank-sum test, is a nonparametric statistical testing method that uses the ranks of values to compare two populations. We used the Mann-Whitney U test throughout this paper to compare Likert scores from two different groups. For comparing more than two populations, we employed the Kruskal-Wallis test, also known as the ANOVA-on-ranks test, which is a nonparametric statistical test for more than two groups. To perform post-hoc power analysis for the aforementioned tests, we used Monte Carlo simulation to estimate the empirical power of Wilcoxon-Mann-Whitney tests and approximated power for the Kruskal-Wallis tests using a chi-square approximation under the null, and a non-central chi-square approximation under the alternative. When gathering Likert data in this work, each experiment was scored on different aspects, resulting in a multi-dimensional Likert data format. To reduce the dimensions when analyzing this data, we used factor analysis based on polychoric correlation. Factor analysis is a statistical method used to describe variability among observed variables based on a lower number of unobserved variables called factors, and polychoric correlation is a statistical method for calculating the correlation between ordinal variables. By applying polychoric correlation during the process of factor analysis, we could extract underlying factors from the multi-dimensional Likert data. All statistical analyses were done using R software version 4.6.0 (R Core Team), using stats package ver.4.6.0, plyr ver.1.8.9, dplyr ver.1.2.1, tidyr ver.1.3.2, ggplot2 ver.4.0.3, ggExtra ver.0.11.0, gridExtra ver.2.3.1, ggpubr ver.1.0.0, likert ver.1.3.5.1, psych ver.2.6.5, and ggforestplotR ver.0.5.0.

##### S2. Code and Data Availability

The data and the code that performs the analysis and generates the figures are available under https://github.com/Ali-esfa/LLM-in-EM. The files *data.RData* and *data.xlsx* in the repository contain the gathered raw data in R format and tabular format, respectively.

#### Supplementary Results

##### S3. Individual Group Analysis

For a deep, granular look into the performance of each LLM, we plotted the Likert scores for every single LLM in each category separately. We utilized bar charts to present the scores, with the height of each bar representing the average Likert score and whiskers marking one standard deviation from the mean (Fig S1). To statistically analyze the differences between LLMs, the Kruskal-Wallis rank sum test was utilized on values in each category to check if there were statistically significant differences between LLM performances. The resulting $p$ values are reported above the corresponding bars on the bar charts (Fig S1A). Observing the simulated doctor visit scores, one might presume some LLMs performed better than some others. For instance, ChatGPT 5.6 Sol, O3, and Claude 4.8 Opus seem to have scored higher than models such as DeepSeek R1 and V4 Pro in most categories, e.g., diagnostic accuracy, treatment plan, and accuracy (Fig S1A-I). However, results of the statistical testing suggest those differences could be due to chance or natural variability of the results, meaning we can't confidently infer one LLM as superior to another in this context. The approximated power of around ~0.12 also concurs with this assessment. The same is true for simulated patient experience results, which show that while there are minor differences between the various LLMs, the difference in the performance is essentially insignificant (Fig 3A-II), with the approximated power of ~0.08.

Resulting scores from the written exam tell a different story, however; LLMs such as ChatGPT 5.6 Sol, O3, and Claude 4.8 Opus demonstrated a clear advantage over some others, for instance, Qwen or DeepSeek R1. The difference in performance between these LLMs is distinctly pronounced in all the categories, with significant differences in diagnosis categories and extremely significant differences in investigation and treatment categories (Fig S1A-III). The approximated power for written exam results was ~0.12.

Performance differences between different human doctor groups, namely different resident groups and specialists, are also plotted as bar charts (Fig S1B). Depending on the experiment and metric, a group scored slightly better than others. Nonetheless, based on the statistical testing outcomes, none of the differences were significant, hinting at virtually the same performance for all the physicians in our experiment, regardless of their experience level.

#### S4. Comparison of This Work with General Benchmarks

Contrasting our results with the general, publicly available benchmarking scores can provide new insights into our measures, validate our approach, and decipher some underlying elements into interpretable factors that influence the performance of LLMs in EM. To that end, we procured benchmarking indices from artificialanalysis.ai and benchlm.ai (Table S1), and plotted them against the factor analysis scores obtained from our benchmarking results (Fig S2). Comparing the general intelligence index with our results, it becomes evident that for LLMs, higher intelligence positively correlates with performance in written exam benchmarking. The reverse of such a relation can be seen with the speed index; higher speed correlates with worse performance in the written exam benchmarks. Interestingly, speed is positively correlated with patient experience, probably because higher speed resulted in smoother conversation with the patient and better comprehension. The scatterplots also show that the more expensive LLMs perform better in the simulated doctor visit and in the diagnosis benchmark from the written exam. By far the most correlated public benchmarking score with our results is the agentic index, which is positively correlated with diagnosis and treatment scores from the written exam, and is significantly correlated with the investigation aspect of the written exam and also the simulated doctor visit score. The multimodal index is also correlated with our scores, but the results are not as dependable since some values in the multimodal index were missing.

Predictive models can also circumvent some steps which are time- and resource-costly. Comparing our sophisticated medicine-specific benchmarking against all-purpose approaches demonstrated the correlation between medical competence and general LLM skill levels, e.g., level of intelligence, speed, reasoning, etc. (Fig S2). The presence of such correlations hints at the existence of a potential machine-learning model that can predict the medical proficiency of an LLM from general information and measures about the LLM. For instance, a predictive model can be trained to estimate the diagnostic skill level using public benchmarks on skills such as general knowledge, medical knowledge, thinking, reasoning, and cognition.

### Supplementary Figures

#### Fig S1. Group Results from Emergency Medicine Benchmarking of AI and Physicians

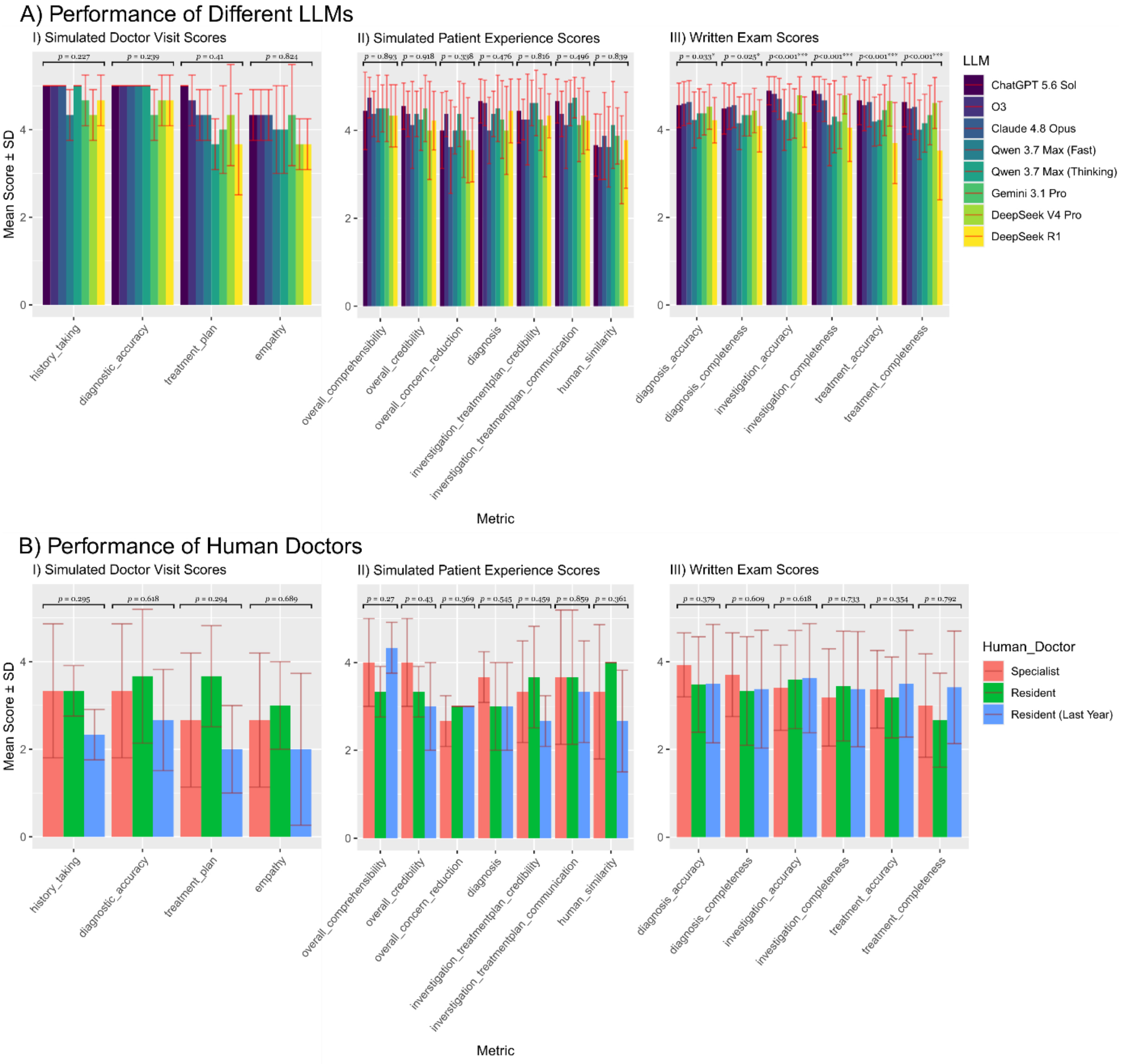


*Figure S1.*

*Results from individual groups of physicians or LLM configurations are plotted as barplots, with the height of each bar representing the average Likert score, whiskers marking one standard deviation from the mean, and p values from Kruskal-Wallis test reported for each group of bars.* ***A.*** *Results of LLMs in three experiment groups of 'Simulated Doctor Visit', 'Simulated Patient Experience', and 'Written Exam', plotted in panels I, II and III, respectively. While LLMs such as ChatGPT, O3, and Claude generally out perform models such as DeepSeek and Qwen, this advantage is statistically significant only in results from the written exam.* ***B.*** *Same as panel A, but representing results from human physicians rather than the LLMs. In general, none of physician groups could significantly outperform others in any of the experiments done in this work.*

#### Fig S2: Our Benchmarking Indices Against Publicly Available Ones

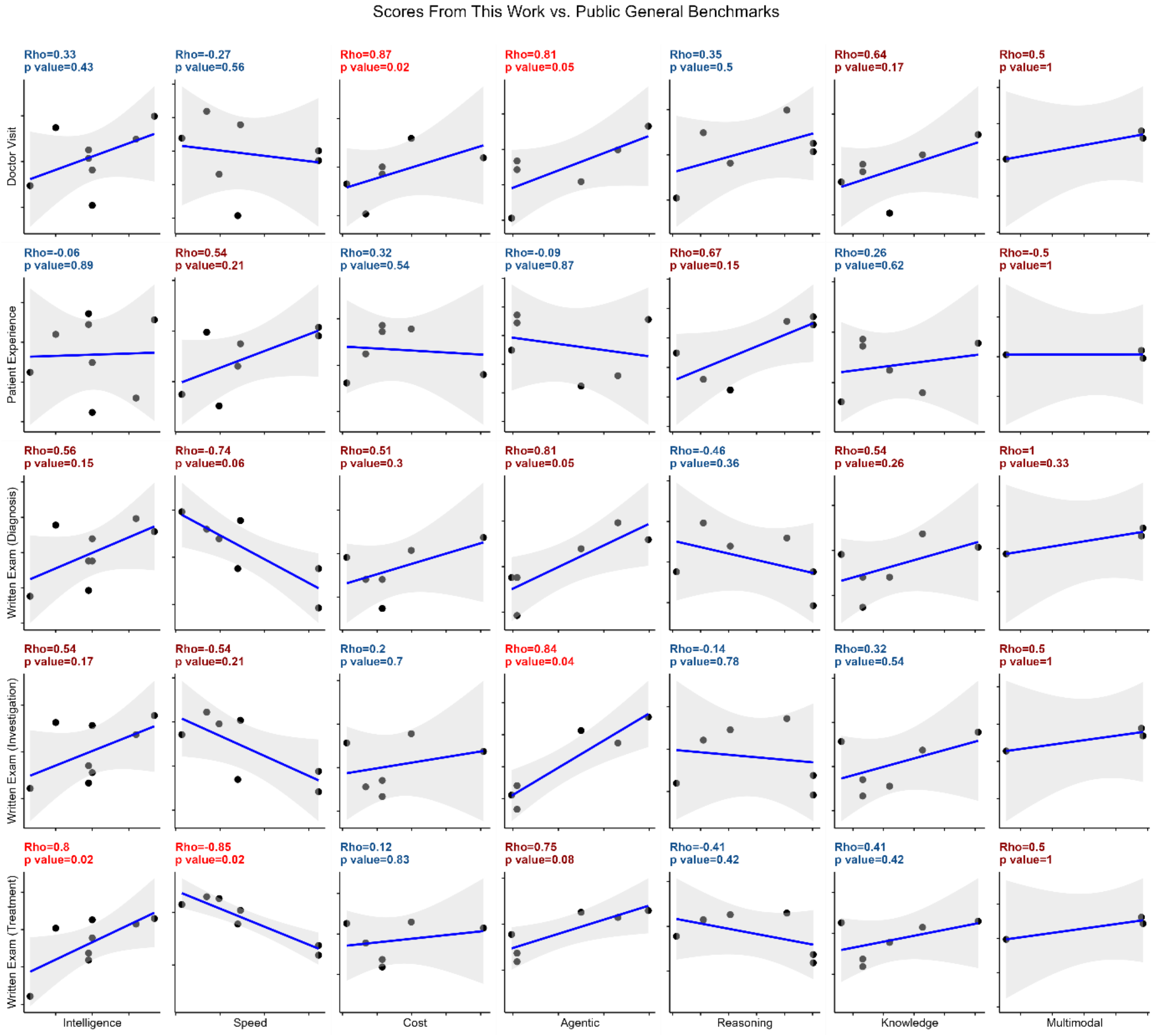


*Figure S2.*

*Pairwise scatterplots are utilized to demonstrate the relationship between different types of scores, with scores from this work plotted in rows (1 factor from the simulated doctor visit, 1 from the simulated patient experience, and 3 from the written exams) and publicly available scores plotted in columns (intelligence, speed, and cost indices obtained from artificialanalysis.ai, and agentic, reasoning, knowledge, and multimodal scores acquired from benchlm.ai). For each scatterplot, Spearman's rank correlation was calculated, and Spearman's ρ (Rho) and p value were reported above each plot using the following color coding: dark red for when the absolute value of the correlation was above 0.5, bright red if, in addition, the p value was equal to or below 0.05, and blue for the rest.*

Fig S3: Factor Analysis and PCA Scree Plots

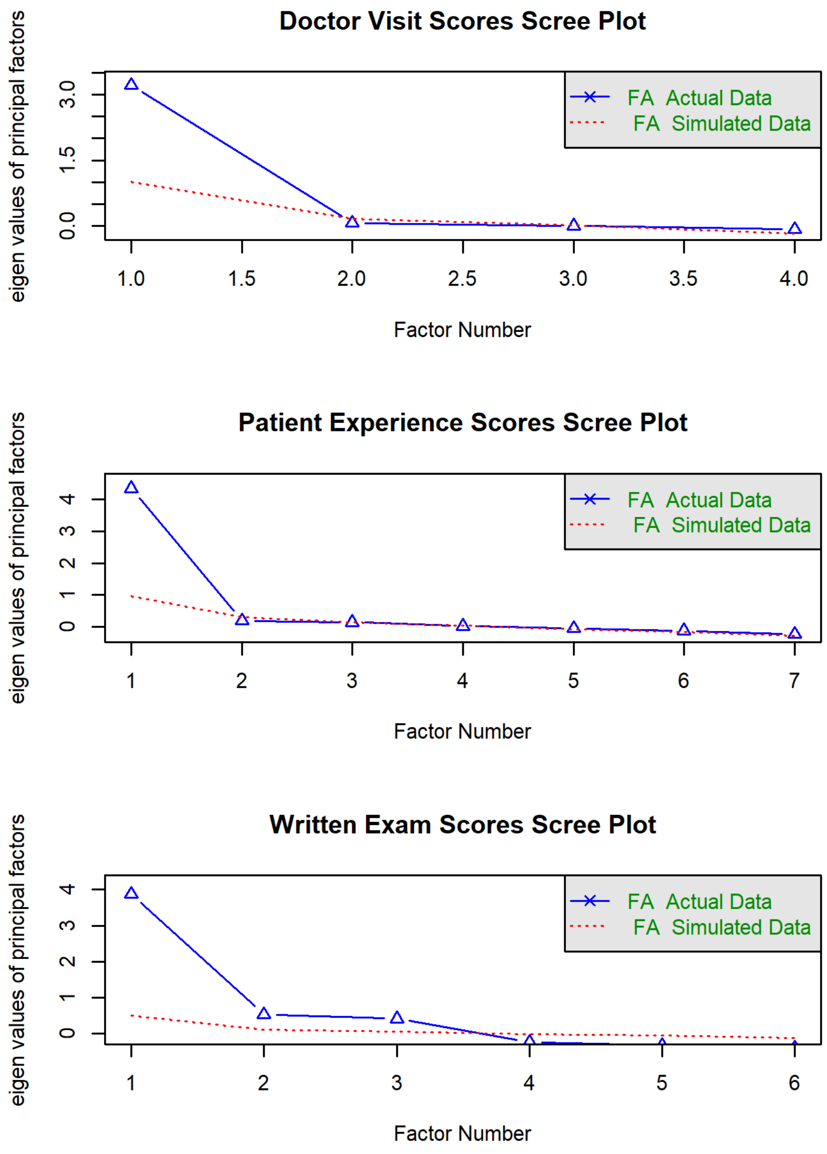


*Figure S3.*

*Scree plots for 'Doctor Visit' (top panel), 'Patient Experience' (middle panel), and 'Written Exam' (bottom panel) acquired data sets. In the top and middle plots, from the second factor onward, eigenvalues are below the random simulation (triangles are below the red dotted line), meaning only one extracted factor represents these data sets properly. However, for the written exam data set, three factors are needed since in the bottom plot, blue triangles go below the red dotted line from the fourth factor onward.*

### Supplementary Tables

Table S1: Public Benchmarking Indices Used for Comparison

| Score | Definition | Unit | Source |
|---|---|---|---|
| Intelligence | Artificial Analysis Intelligence Index v4.3.2, comprising 10 different evaluating benchmarks | arbitrary unit | https://artificialanalysis.ai |
| Speed | LLM output speed | tokens per second | https://artificialanalysis.ai |
| Cost | Cost per Intelligence Index task | Dollars | https://artificialanalysis.ai |
| Agentic | Measure of tool use, browser research, and computer-use workflows | arbitrary unit | https://benchlm.ai/ |
| Reasoning | Measure of Logical reasoning and problem solving | arbitrary unit | https://benchlm.ai/ |
| Knowledge | Measure of General knowledge and factual understanding | arbitrary unit | https://benchlm.ai/ |
| Multimodal | Measure of Vision, document, and grounded enterprise workflow benchmarks | arbitrary unit | https://benchlm.ai/ |
